# T-cell dysregulation is implicated in the encephalopathy of prematurity

**DOI:** 10.64898/2026.10.15.26365621

**Authors:** Rebekah Smikle-Bernard, Manuel Blesa Cábez, Kadi Vaher, Ray Amir, Katie Mckinnon, Lee Murphy, Michael J. Thrippleton, Alan J. Quigley, Mark E. Bastin, Rebecca M. Reynolds, Heather C. Whalley, Riccardo E. Marioni, Simon R. Cox, James P. Boardman

**Affiliations:** Centre for Reproductive Health, Institute for Regeneration and Repair, University of Edinburgh, Edinburgh, EH16 4UU, UK; Institute for Neuroscience and Cardiovascular Research, University of Edinburgh, Edinburgh, EH16 4TJ, UK; Edinburgh Clinical Research Facility, University of Edinburgh, Western General Hospital, Edinburgh, EH4 2XU, UK; Edinburgh Imaging Facility, Royal Infirmary of Edinburgh, Edinburgh, EH16 4TJ, UK; Department of Radiology, Royal Hospital for Children and Young People, Edinburgh, EH16 4TJ, UK; Lothian Birth Cohorts, Department of Psychology, University of Edinburgh, Edinburgh, EH8 9JZ, UK; Centre for Cardiovascular Science, Queen’s Medical Research Institute, University of Edinburgh, Edinburgh, EH16 4TJ, UK; Institute of Genetics and Cancer, University of Edinburgh, Edinburgh, EH4 2XU, UK

**Keywords:** T cells, CRTAM, epigenetics, MRI, preterm birth, white matter

## Abstract

Preterm birth is strongly linked to systemic inflammation and encephalopathy of prematurity, which underlies subsequent cognitive impairment. T-cell immunity contributes to brain-immune crosstalk across the life course but its role in the pathogenesis of encephalopathy of prematurity is unknown. Protein epigenetic scores are DNA methylation (DNAm)-based proxies of circulating proteins. We tested the hypothesis that T-cell dysregulation has a specific role in child neurodevelopment following preterm birth.

Multimodal data were collected from 144 preterm infants (median gestational age at birth 29^+5^ weeks [range 23^+3^ to 32^+6^]) recruited to a prospective cohort study: saliva DNAm for protein epigenetic score derivation and brain MRI, both at term-equivalent age, and the Mullen Scales of Early Learning Composite score at 5 years from a subset. We investigated associations of the epigenetic scores for cytotoxic and regulatory T cell-associated molecule (DNAm CRTAM) and C-reactive protein (DNAm CRP; a non-specific marker of inflammation) with MRI metrics of encephalopathy of prematurity: global and regional brain volumes, and general factors of diffusion tensor imaging and neurite orientation dispersion and density imaging metrics averaged across 42 white matter tracts. We tested whether DNAm CRTAM explains more variance than DNAm CRP in MRI metrics of the encephalopathy of prematurity and cognitive scores in childhood.

Higher neonatal DNAm CRTAM was associated with lower general factor of fractional anisotropy (β = −0.252, *q* = 0.011) and higher general factors of radial diffusivity (β = 0.215, *q* = 0.039), mean diffusivity (β = 0.198, *q* = 0.047) and orientation dispersion index (β = 0.201, *q* = 0.048), independent of inflammatory morbidities and other biological covariates after false discovery rate correction. DNAm CRTAM explained more variance than DNAm CRP (maximum incremental R^2^ = 4.2% vs 1.5%) in diffusion MRI metrics. Higher neonatal DNAm CRTAM was associated with lower cognitive scores at 5 years, independent of DNAm CRP and other covariates (β = −0.332, *P* = 0.008), and explained more variance in cognitive scores than DNAm CRP (incremental R^2^ = 9.4% vs 1.2%).

An epigenetic proxy of CRTAM in preterm infants is associated with both encephalopathy of prematurity in the neonatal period and lower cognitive performance at 5 years, explaining greater incremental variance than a non-specific marker of inflammation. The data implicate T-cell dysregulation in the pathophysiology of atypical neurodevelopment following preterm birth.

## Introduction

Preterm birth affects 9.9% of live births globally, which equates to approximately 13.4 million infants per year.^1^ Infants born preterm are at an increased risk of cognitive difficulties that persist across the life course, with no effective treatments currently available to improve outcomes.^2^ The neural substrate underlying long-term cognitive impairment includes macro- and microstructural alterations in the developing brain, collectively termed the encephalopathy of prematurity (EoP).^3,4^ Elucidating the biological axes linking preterm birth with EoP is a priority for understanding the pathogenesis of EoP and developing targeted treatments.

MRI-based macrostructural features of EoP are reduced global and regional brain volumes and altered cortical morphology, accompanied by increased volume of intraventricular and extra-axial fluid compartments.^5–8^ The microstructural signature of EoP in preterm infants at term-equivalent age is diffuse white matter dysmaturation inferred by decreased fractional anisotropy (FA) and neurite density index (NDI), and increased mean and radial diffusivity (MD, RD), detected using diffusion tensor imaging (DTI) and neurite orientation dispersion and density imaging (NODDI).^9–12^ To capture the generalised nature of white matter dysconnectivity across major tracts that is characteristic of EoP, as well as reduce dimensionality of highly correlated diffusion metrics across several tracts, we have previously derived general (*g*-)factors of white matter microstructure from neonatal diffusion MRI data (e.g., *g*FA, *g*MD, *g*RD, *g*NDI).^13,14^ MRI features of EoP are associated with later neurocognitive outcomes and so serve as intermediate phenotypes for investigating upstream determinants^15–17,18–21^

Systemic immune dysregulation is a potentially modifiable factor linking preterm birth with atypical brain development.^22,23^ Inflammatory comorbidities of preterm birth such as histologic chorioamnionitis, sepsis, bronchopulmonary dysplasia and necrotising enterocolitis are associated with differences in white matter microstructure.^24–27^ Elevated inflammatory blood proteins in preterm infants are also linked to abnormal white matter development on MRI^22,28^ and later childhood cognitive and behavioural outcomes^29,30^, but previous technologies and study designs leave uncertainties about which immune axes mediate the link between preterm birth and child development.

Recent studies draw attention to T-cell dysregulation in brain-immune crosstalk across the life course. Autism is associated with altered effector T cell phenotypes (CD4+, CD8+), lower proportions of T regulatory cells (T_regs_), and altered activation of T cells.^31^ Preterm infants show similarly skewed helper subsets and differences in receptor expression and functional characteristics of T_regs_.^32^ Cytotoxic and regulatory T cell-associated molecule (CRTAM), also known as class-I Major Histocompatibility Complex restricted T cell-associated molecule, is highly expressed in activated CD4+, CD8+ and Natural Killer T cells and in cerebellar Purkinje neurons.^33,34^ *CRTAM* is an Alzheimer’s disease risk gene^35^ and CRTAM is upregulated in plasma and cerebrospinal fluid of adults with diagnosed and prodromal Alzheimer’s disease compared to age-matched controls.^36,37^ Higher plasma CRTAM is associated with lower processing speed in adults,^38^ and it contributes to a proteomic signature of global cognitive function in adults with metabolic morbidities.^39^

Circulating proteins can be proxied using DNA methylation (DNAm) to derive protein ‘epigenetic scores’ (EpiScores).^40^ Protein EpiScores offer a more stable means of profiling chronic inflammation, showing higher test-retest reliability and stronger associations with disease outcomes than their protein counterparts.^41–44^ The EpiScore of C-reactive protein (DNAm CRP), calculated using neonatal salivary DNAm is associated with low gestational age, perinatal inflammatory morbidities and differences in brain volume and white matter microstructure in preterm neonates (key hallmarks of EoP on MRI^17,45^) and with childhood cognition.^46^ However, although CRP, being an acute phase protein, is relatively sensitive as a marker of systemic inflammation,^47^ it lacks specificity so does not provide insight about immune pathways. The neonatal EpiScore of CRTAM (DNAm CRTAM) is higher in preterm infants compared to term-born infants,^45^ and is associated with lower cognitive performance at 5 years of age.^46^ DNAm CRTAM has also been shown to associate with incidence of Alzheimer’s disease up to 8 years before diagnosis^40^. In previous work, there was no association between neonatal DNAm CRTAM and perinatal inflammatory burden,^46^ which suggests a potential role of T-cell pathway signalling distinct from acute inflammatory processes.^48^

To elucidate the specific role of T-cell pathways in child development after preterm birth, we studied a longitudinal cohort of children born at less than 33 weeks’ gestation. We tested associations between DNAm CRTAM and structural and diffusion MRI features of EoP, and investigated whether DNAm CRTAM explains more variance than DNAm CRP, a non-specific marker of acute inflammation, in MRI features of EoP and 5-year cognitive performance.

## Materials and methods

### Study population

Participants were 144 preterm children (born at <33 weeks’ gestation) with both neonatal DNAm data and MRI acquisition. Participants were recruited into the Theirworld Edinburgh Birth Cohort, a prospective, longitudinal study designed to investigate the impact of preterm birth on brain development and outcomes.^49^ Cohort exclusion criteria were major congenital malformations, chromosomal abnormalities, congenital infection, parenchymal brain injury (cystic periventricular leukomalacia, haemorrhagic parenchymal infarction or post - haemorrhagic ventricular dilation).^50^ Ethics approval was obtained from the National Research Ethics Service, South East Scotland Research Ethics Committee (13/SS/0143) and NHS Lothian Research and Development (16/SS/0154). Parents provided written informed consent in accordance with the Declaration of Helsinki. The STROBE guidelines were used for reporting of this observational study. Blinding and randomisation were not performed because the study did not involve allocation to intervention or control groups.

### Clinical and demographic measures

Clinical and demographic data were collected through questionnaires administered to primary caregivers and extraction from medical records. We included the following perinatal inflammatory comorbidities: histologic chorioamnionitis was defined as presence of inflammatory response in placental membranes of any grade or stage; sepsis was defined as blood culture positive for pathogenic organism (culture positive) or clinically suspected infection treated with intravenous antibiotics for ≥5 days (culture negative); necrotising enterocolitis was defined as medical treatment for ≥7 days or surgical treatment; bronchopulmonary dysplasia was defined as need for supplemental oxygen therapy or respiratory support at 36 weeks corrected gestational age. Comorbidities were coded as binary variables (present/absent). Birthweight z-scores were calculated according to INTERGROWTH-21st standards.^51^ Maternal education, an individual-level indicator of socioeconomic status, was coded as a binary variable of university undergraduate degree or higher (Yes/No).

### DNA methylation and EpiScore calculation

Saliva samples for DNAm were obtained at term-equivalent age. Saliva was collected in Oragene OG-575 Assisted Collection kits, by DNA Genotek, and DNA extracted using prepIT.L2P reagent (DNA Genotek, ON, Canada).

Details of methylation measurement have been described previously.^45^ Briefly, DNA was bisulphite converted and methylation levels were measured using Illumina HumanMethylationEPIC BeadChip (Illumina, San Diego, CA, USA) at the Edinburgh Clinical Research Facility (Edinburgh, UK). Arrays were imaged on the Illumina iScan or HiScan platform, and genotypes called automatically using GenomeStudio Analysis software 2011.1 (Illumina). DNAm was processed in four batches, and so processing batch was included as a categorical covariate in downstream analyses, as previously described.^46^

Details of DNAm data pre-processing have been outlined previously.^45,46^

For each participant, protein EpiScores were obtained by multiplying the methylation proportion at a given CpG by the effect size from previous studies where protein levels were compared with DNAm patterns using epigenome-wide association studies (EWAS) or elastic net regression.^40,45^ The CRTAM EpiScore used was derived using predictor weights calculated from elastic net penalised regression models reported by Gadd and colleagues,^40^ where DNAm CRTAM (based on 64 CpG sites) explained ∼10% of variance in protein levels in independent tests. The original training dataset for DNAm CRTAM was the Scottish Lothian Birth Cohort 1936 (LBC1936^52^; n = 875, mean age 73 years), where DNAm was quantified in blood using HumanMethylation450K BeadChip. Independent validation datasets included a subset of Generation Scotland:STRADL^53^ (n = 778, mean age 66 years) and the LBC1921^52^ (n = 162, mean age 87 years). The CRP EpiScore weights were from the seven-CpG variation described by Barker and colleagues,^54^ based on the CRP EpiScore originally calculated by Ligthart and colleagues, where DNAm CRP explained ∼4% of variance in protein levels.^55^ Ligthart et al.’s EWAS training dataset included 9 European adult cohorts (n = 8,863, mean ages 60-87 years) and a validation dataset of 4 African American cohorts (n = 4,111, mean ages 41-73 years), using the HumanMethylation450K BeadChip for DNAm quantification in whole blood.

Ligthart et al.’s CRP EpiScore was previously shown to correlate with inflammation-related conditions, including perinatal inflammatory exposures and adult chronic morbidities, as well as brain development and cognitive outcomes in preterm and general paediatric cohorts.^17,46,54,55^ This seven-CpG variation also demonstrates stronger associations with serum CRP at birth than the more recently developed CRP EpiScore integrating 1,511 CpG sites.^56^

### MRI acquisition

Structural and diffusion MRI data were acquired at Edinburgh Imaging Facility, Royal Infirmary of Edinburgh, University of Edinburgh, UK, using a MAGNETOM Prisma 3T MRI clinical scanner (Siemens Healthcare GmbH, Erlangen, Germany) and 16-channel phased-array paediatric head and neck coil. All infants were scanned around term-equivalent age, according to the published protocol.^49^

3D T2-weighted (T_2_w) SPACE images (TR = 3200 ms, TE = 409 ms) with 1 mm isotropic voxels were acquired. Diffusion MRI data were acquired in two separate acquisitions to reduce the time needed to reacquire any data lost to motion artefacts: the first consisted of eight baseline volumes [*b* = 0 s/mm2 (*b*0)] and 64 volumes with *b* = 750 s/mm^2^; the second consisted of 8 *b*0, 3 volumes with *b* = 200 s/mm^2^, 6 volumes with *b* = 500 s/mm^2^ and 64 volumes with *b* = 2500 s/mm^2^. An optimal angular coverage for the sampling scheme was applied.^57^ In addition, an acquisition of three b0 volumes with an inverse phase encoding direction was performed. Diffusion MRI volumes were acquired using single-shot spin-echo EPI with 2-fold simultaneous multi-slice and 2-fold in-plane parallel imaging acceleration and 2 mm isotropic voxels; both diffusion acquisitions had the same parameters (TR = 3500 ms, TE = 78.0 ms). Images affected by motion artefact were reacquired multiple times as required; diffusion MRI acquisitions were repeated if the signal loss was seen in three or more volumes.

Infants were fed and wrapped and allowed to sleep naturally in the scanner. Pulse oximetry, electrocardiography and temperature were monitored. Flexible earplugs and neonatal earmuffs (MiniMuffs, Natus) were used for acoustic protection. Scans were supervised by a doctor or nurse trained in neonatal resuscitation. Structural images were reported by an experienced paediatric radiologist.

### Diffusion MRI preprocessing

Details on diffusion MRI pre-processing have been previously outlined.^58^

In brief, the two diffusion MRI acquisitions were first concatenated before preprocessing, then denoised using a Marchenko-Pastur-PCA-based algorithm^59–61;^ eddy current, head movement and EPI geometric distortions were corrected using outlier replacement and slice-to-volume registration^62–66;^ bias field inhomogeneity correction was performed by calculating bias field of mean b0 volume and applying the correction to all volumes.^67^

### MRI features of encephalopathy of prematurity

#### Global and regional brain volumes

T_2_w images were processed using the Developing Human Connectome Project pipeline to obtain bias field-corrected T_2_w and the brain mask and tissue segmentation.^68^ Volumes extracted included cortical grey matter, deep grey matter, white matter, hippocampi and amygdalae, cerebellum, brainstem, cerebrospinal fluid (CSF) and ventricles.

#### Tract segmentation and extraction of tract-averaged diffusion MRI metrics for calculation of general factors of white matter microstructure

From the diffusion MRI data we calculated the DTI maps (FA, MD, AD, RD), the neurite density index (NDI) map using Spherical Mean Technique^69,70^ and the orientation dispersion index (ODI) map from NODDI. DTI maps were calculated using only the *b* = 750 s/mm^2^ shell. ODI maps were computed with Accelerated Microstructure Imaging via Complex Optimisation, AMICO.^71^

A total of 42 tracts were generated using XTRACT.^72,73^ The tracts were thresholded to 0.005 and then averaged in template space. Finally, tracts were binarised keeping only voxels common to 30% of the subjects and mapped back to subjects’ native space.

Representative brain maps for tissue segmentation, DTI metrics and NODDI metrics, as well as tract segmentation are provided in Fig. 1.

**Figure 1.**
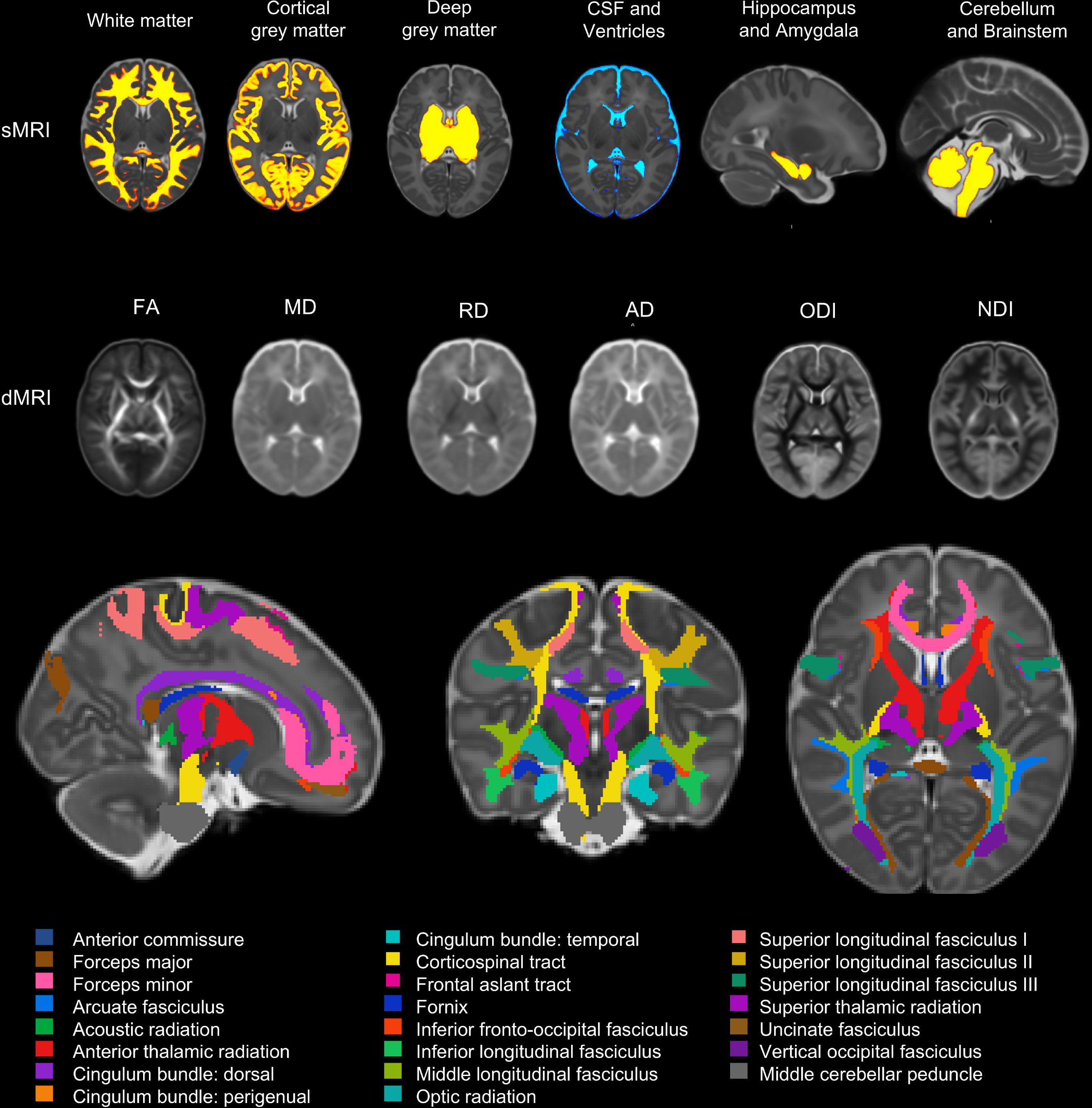
Representative brain maps and reconstructed white matter tracts. Top panel: Segmentation of the brain tissues of interest from structural MRI (sMRI), overlaid on the Developing Human Connectome Project 40-week T_2_w template. Fluid compartments and hindbrain tissue are grouped for visualisation. Middle panel: Diffusion MRI (dMRI) maps including diffusion tensor imaging maps and neurite orientation dispersion and density imaging maps. dMRI maps are averaged over participants with complete sMRI and dMRI acquisitions (n = 132). Bottom panel: Orthogonal sagittal (left), coronal (centre) and axial (right) views showing the XTRACT-derived white matter tracts overlaid on the Developing Human Connectome Project 40-week T_2_w template. The three views correspond to the same three-dimensional anatomical location and were selected to provide broad visualisation of the reconstructed tracts. CSF, cerebrospinal fluid; FA, fractional anisotropy; MD, mean diffusivity; RD, radial diffusivity; AD, axial diffusivity; ODI, orientation dispersion index; NDI, neurite density index.

### Cognitive assessment

At 5 years of age, 67 children were assessed using the Mullen Scales of Early Learning (MSEL).^74^ MSEL Early Learning Composite provides a measure of general cognitive performance derived from four subscales, namely visual reception, fine motor, receptive language and expressive language. The Early Learning Composite is a standard score with normative mean of 100 and SD of 15.

### Statistical analysis

Statistical analyses were performed in R (version 4.5.1).^75^ Relevant packages are detailed below.

#### Participant characteristics

Maternal and infant clinical and demographic characteristics were summarised using descriptive statistics. Continuous variables were assessed for normality using the Shapiro-Wilk test. Normally distributed continuous variables are presented as mean and standard deviation, while non-normally distributed continuous variables are presented as median and range. Categorical variables are presented as frequencies and percentages.

#### Calculation of *g* factors for each diffusion MRI metric

Calculation of single-metric *g* factors from neonatal diffusion MRI data has been previously described.^14^ In the current study, principal component analysis was conducted separately for each metric (FA, MD, RD, AD, NDI, ODI) across 42 tracts to quantify the proportion of shared variance between them; principal component analysis was implemented using the “psych” package in R (cran.r-project.org/web/packages/psych/psych.pdf). Each subject was described by 42 features, computed as the tract-averaged values of each metric across each tract (Supplementary Fig. 1). For each metric, the first unrotated principal component scores were extracted as the *g* factor (e.g. *g*FA). Scree plots showing the proportion of explained variance and eigenvalues for the first ten principal components per diffusion MRI metric are illustrated in Supplementary Fig. 2. Tract loadings and proportion of explained variance for each derived *g* factor are presented in Supplementary Table 1.

#### Sensitivity power analysis

To evaluate the power afforded by the sample size (N = 144), we used the *pwr.f2.test* implemented using the “pwr” package in R (cran.r-project.org/web/packages/pwr/pwr.pdf). We accounted for the main predictor (DNAm CRTAM) and the five covariates included in baseline models. Using standard linear approximations, we estimated the minimum detectable effect size (R^2^) at α = 0.05 with 80% power. Sensitivity power analysis indicated that the study was sufficiently powered to detect small-to-moderate effects (R^2^ = 5.4%).

#### DNAm CRTAM and neonatal brain structure associations

To examine associations of DNAm CRTAM and preterm neonatal brain structure, data were analysed using linear regression. Outcome variables were EoP-relevant brain MRI metrics. Covariates in baseline models included: gestational age at birth, as it is associated with DNAm CRTAM^45^ and with EoP-relevant MRI features in a dose-dependent manner^7,76^; gestational age at DNAm sampling/MRI scan; infant sex, given established sex differences in neonatal tissue DNA methylation and in neonatal brain structure^77,78^; birthweight Z-score, an estimate of fetal growth restriction, which is associated with antenatal chronic low-grade inflammation and brain structure^79,80^; and DNAm processing batch. In sensitivity analyses, inflammation-adjusted models additionally included histologic chorioamnionitis, bronchopulmonary dysplasia, necrotising enterocolitis and sepsis simultaneously, to assess whether DNAm CRTAM explained variance in MRI metrics beyond clinical inflammatory exposures. Complete case analyses were conducted for each model. Performance of regression models was assessed using the “performance” package in R (cran.r-project.org/web/packages/performance/performance.pdf). Visual inspection of diagnostic plots suggested no regression assumptions were violated (example for fully adjusted model in Supplementary Fig. 3).

Incremental R^2^ was used to quantify the additional variance in each MRI metric explained by DNAm CRTAM beyond covariates. For the baseline model, incremental R^2^ was calculated as the difference in unadjusted R^2^ between the model including DNAm CRTAM and baseline covariates, and the corresponding covariate-only model. For the inflammation-adjusted model, incremental R^2^ was calculated as the difference in unadjusted R^2^ between the fully adjusted model including DNAm CRTAM and the corresponding fully adjusted model without DNAm CRTAM.

For MRI metrics with significant associations, we examined whether there is an interaction between DNAm CRTAM and gestational age at birth by including a product interaction term (DNAm CRTAM x gestational age at birth) in baseline models.

Prior to analyses, global MRI volumetric measures were regressed on intracranial volume to remove variance related to head size, avoid multicollinearity with other biological covariates, and produce brain-specific measures usable consistently across hierarchical models. Residuals from these linear regression models were taken forward for downstream analyses.

Continuous variables were scaled to have a mean of zero and variance of one. All statistical tests were two-tailed tests, with a nominal significance threshold of α = 0.05. *P*-values were corrected for multiple comparisons using the false discovery rate (FDR) method (Benjamini-Hochberg procedure),^81^ with significance defined as *q* <0.05. FDR correction was applied within imaging modality, that is, separately for volumetric and diffusion MRI features, to reflect their distinct biological interpretations. Given the moderate to high correlations among neuroimaging features, nominal uncorrected associations at *P* <0.05 are reported alongside FDR-adjusted results.

#### Unique contribution of DNAm CRTAM to neonatal brain structure beyond DNAm CRP

The correlation between DNAm CRTAM and DNAm CRP was assessed using Pearson’s correlation. Given previous work demonstrating associations between DNAm CRP and EoP-relevant imaging features,^17^ we investigated whether DNAm CRTAM explains more incremental variance in MRI features than DNAm CRP. MRI metrics showing significant associations were taken forward into secondary models which additionally covaried for DNAm CRP. We examined change in coefficient of DNAm CRTAM after adjusting for DNAm CRP. Williams’ test for differences in dependent correlations^82^ was also used to compare association magnitudes of both EpiScores, treating standardised betas as correlations for this purpose and accounting for the predictors’ intercorrelations. Williams’ test (r.test) was implemented using the “psych” package in R (cran.r-project.org/web/packages/psych/psych.pdf). We also calculated incremental R^2^ of models with and without each EpiScore, to evaluate the added explanatory value of DNAm CRTAM or DNAm CRP beyond the other EpiScore and other covariates.

#### Unique contribution of DNAm CRTAM to childhood cognition beyond DNAm CRP

We investigated whether DNAm CRTAM explains more incremental variance in 5-year Early Learning Composite than DNAm CRP using multiple linear regression. We calculated incremental R^2^ of models with and without each EpiScore, to evaluate the added explanatory value of DNAm CRTAM or DNAm CRP beyond the other EpiScore and other covariates.

In a separate model where MSEL Early Learning Composite was regressed on DNAm CRTAM, we included a product interaction term (DNAm CRTAM x gestational age at birth) to additionally assess whether the association is dependent on the degree of prematurity.

Covariates in both analyses were selected based on their potential to act as confounders or because their inclusion could improve precision of model estimates: gestational age at sampling, infant sex and DNAm processing batch.

## Results

### Participant characteristics

The study group consisted of 144 preterm infants; demographic and clinical characteristics of whom are provided in Table 1. Forty-three (31.9%) infants had histologic chorioamnionitis, 31 (21.5%) had an episode of postnatal sepsis, 35 (24.6%) had bronchopulmonary dysplasia and 6 (4.2%) developed necrotising enterocolitis.

**Table 1.** Demographic and clinical characteristics of participants.

| Characteristics | N = 144 |
| --- | --- |
| Sex, n (%) |  |
| Female | 61 (42) |
| Male | 83 (58) |
| Gestational age at birth / weeks + days, median (range) | 29 +5 (23 +3, 32 +6) |
| Gestational age at DNAm sample and MRI scan / weeks + day, mean (SD) | 40 +5 (1.53) |
| Birthweight / g, median (range) | 1291 (500, 2510) |
| Birthweight z-score, median (range) | 0.16 (-3.13, 2.14) |
| Histological chorioamnionitis, n (%) | 43/135 (31.9) |
| Sepsis, n (%) | 31/144 (21.5) |
| Bronchopulmonary dysplasia, n (%) | 35/142 (24.6) |
| Necrotising enterocolitis, n (%) | 6/144 (4.2) |
| Maternal age / years, mean (SD) | 31.99 (5.71) |
| Maternal body mass index / median (range) | 25.45 (16.40, 48.00) |
| Smoked during pregnancy, n (%) | 23/141 (16.3) |
| Antenatal corticosteroid exposure, n (%) | 137 (95.1) |
| Antenatal MgSO<sub>4</sub> for fetal neuroprotection, n (%) | 112 (77.8) |
| Mother university degree, n (%) | 71 (49.3) |

Normally distributed continuous variables are presented as mean and standard deviation (SD), while non-normally distributed continuous variables are presented as median and range. Categorical variables are presented as frequencies and percentages. For inflammatory morbidities (histologic chorioamnionitis, sepsis, bronchopulmonary dysplasia and necrotising enterocolitis) and smoking during pregnancy, fractions are presented to account for missing data. DNAm = DNA methylation, MgSO<sub>4</sub> = Magnesium sulphate.

### Associations between DNAm CRTAM and MRI features of the encephalopathy of prematurity

#### i) Diffusion MRI

In baseline models adjusted for gestational age at scan, gestational age at birth, infant sex and DNAm processing batch, higher DNAm CRTAM was significantly associated with lower *g*FA (β = −0.239 [95% CI −0.396, −0.083], *q* = 0.018; incremental R^2^ = 4.6%). Nominally significant positive associations were also observed in baseline models of *g*RD (β = 0.184 [95% CI 0.010, 0.357], *P* = 0.038, *q* = 0.077; incremental R^2^ = 2.7%) and *g*ODI (β = 0.206 [95% CI 0.021, 0.391], *P* = 0.029, *q* = 0.077; incremental R^2^ = 3.4%), (Fig. 2A; Supplementary Table 2).. Associations remained significant when inflammatory morbidities were accounted for: *g*FA (β = −0.252, *q* = 0.011), *g*RD (β = 0.215, *q* = 0.039), *g*MD (β = 0.198, *P* = 0.024, *q* = 0.047), *g*ODI (β = 0.201, *P* = 0.032, *q* = 0.048), with DNAm CRTAM explaining 3.2-5.1% incremental variance above comorbidities and other covariates. DNAm CRTAM therefore contributes to variance explained in global white matter integrity beyond the effect of clinical exposures. There was no evidence of an interaction between gestational age at birth and DNAm CRTAM in relation to any diffusion MRI metric (Supplementary Table 3).

**Figure 2.**
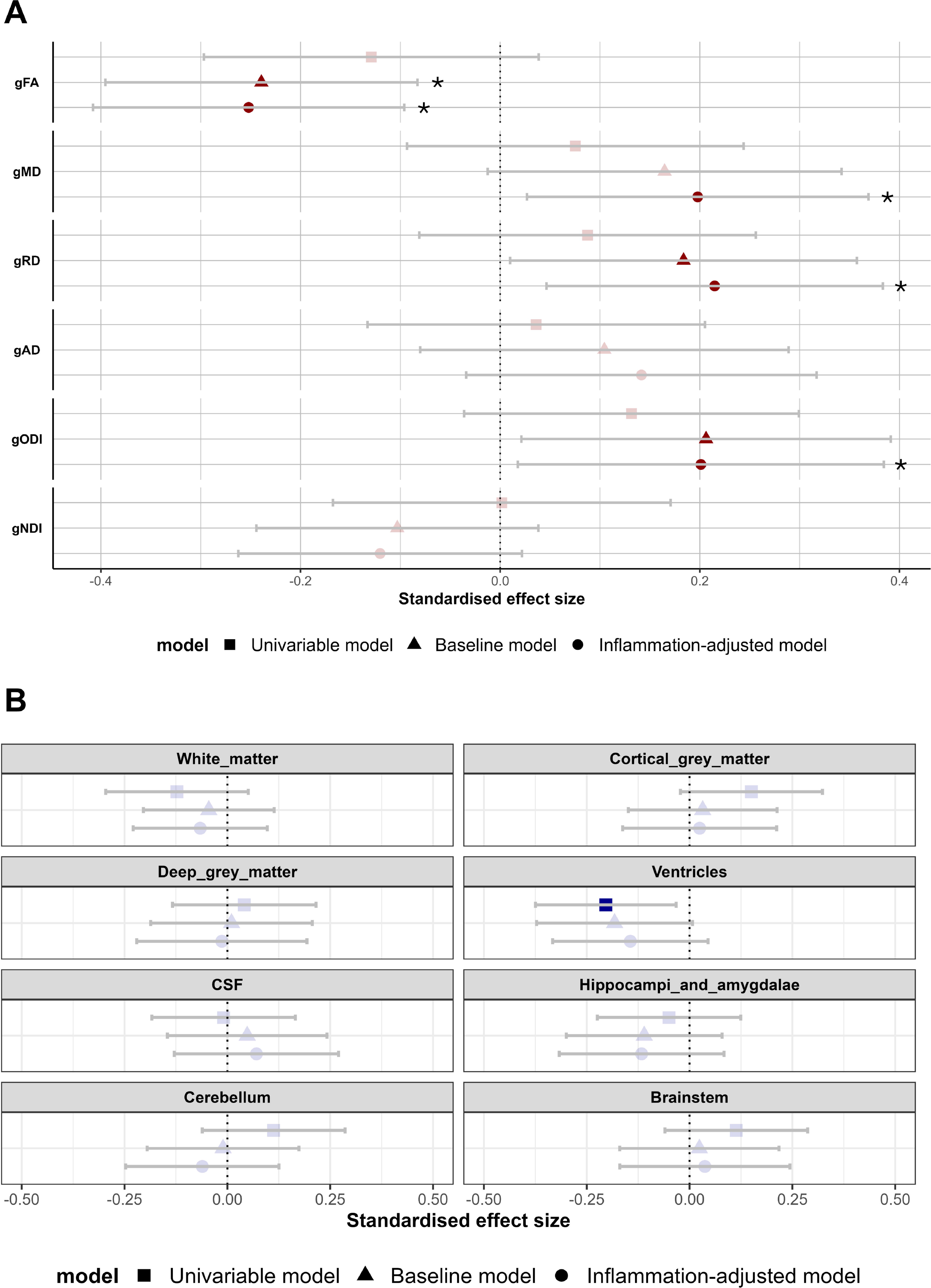
Associations of DNAm CRTAM with MRI features of the encephalopathy of prematurity. Point shapes show standardised regression coefficients with 95% confidence intervals for associations of DNAm CRTAM with A) general factors of diffusion MRI metrics (n = 139) and B) global and regional brain volumes (n = 137). Significant associations at the nominal threshold *P* <0.05 are denoted by dark filled shapes. Asterisks indicate statistically significant associations at *q* <0.05. Baseline models control for gestational age at birth, gestational age at scan/sampling, infant sex, birthweight Z-score and DNAm processing batch. Inflammation-adjusted models covary for inflammatory morbidities simultaneously: histologic chorioamnionitis, sepsis, bronchopulmonary dysplasia and necrotising enterocolitis. g, general factor; FA, fractional anisotropy; MD, mean diffusivity; RD, radial diffusivity; AD, axial diffusivity; ODI, orientation dispersion index; NDI, neurite density index; CSF, cerebrospinal fluid.

#### ii) Structural MRI

Higher DNAm CRTAM was nominally associated with smaller ventricular volume in univariable models (−0.204 [95% CI −0.375, −0.033], *P* = 0.02, *q* = 0.160), but this association did not survive correction for multiple comparisons or adjustment for other covariates. No other volumetric associations were observed (Fig. 2B; Supplementary Table 2).

### Unique contributions of DNAm CRTAM to variance in preterm neonatal brain structure beyond DNAm CRP

DNAm CRTAM and DNAm CRP were weakly correlated (*r* = 0.21, *P =* 0.013; Supplementary Fig. 4), suggesting that DNAm CRTAM reflects an immune-related signal that is partly distinct from that captured by DNAm CRP. Table 2 reports associations between DNAm CRTAM and *g*FA, *g*RD and *g*ODI, after adjustment for DNAm CRP. DNAm CRTAM explained approximately 4.2%, 2.4% and 2.5% incremental variance in each metric, respectively. By comparison, DNAm CRP explained no more than 1.5% incremental variance in these metrics. Effect size estimates of DNAm CRTAM were modestly attenuated after adjustment (decreases in β ranging 0.006 to 0.025), suggesting that some variance captured by DNAm CRTAM overlaps with that of DNAm CRP. However, DNAm CRTAM remained statistically significantly associated with *g*FA (β = −0.233, *P* = 0.004, *q* = 0.027). Across other MRI metrics of EoP, a modality-specific pattern was observed: DNAm CRTAM consistently explained more incremental variance in global diffusion metrics of white matter microstructure, whereas DNAm CRP tended to explain more incremental variance in global and regional brain volumes (Supplementary Table 3).

**Table 2.**
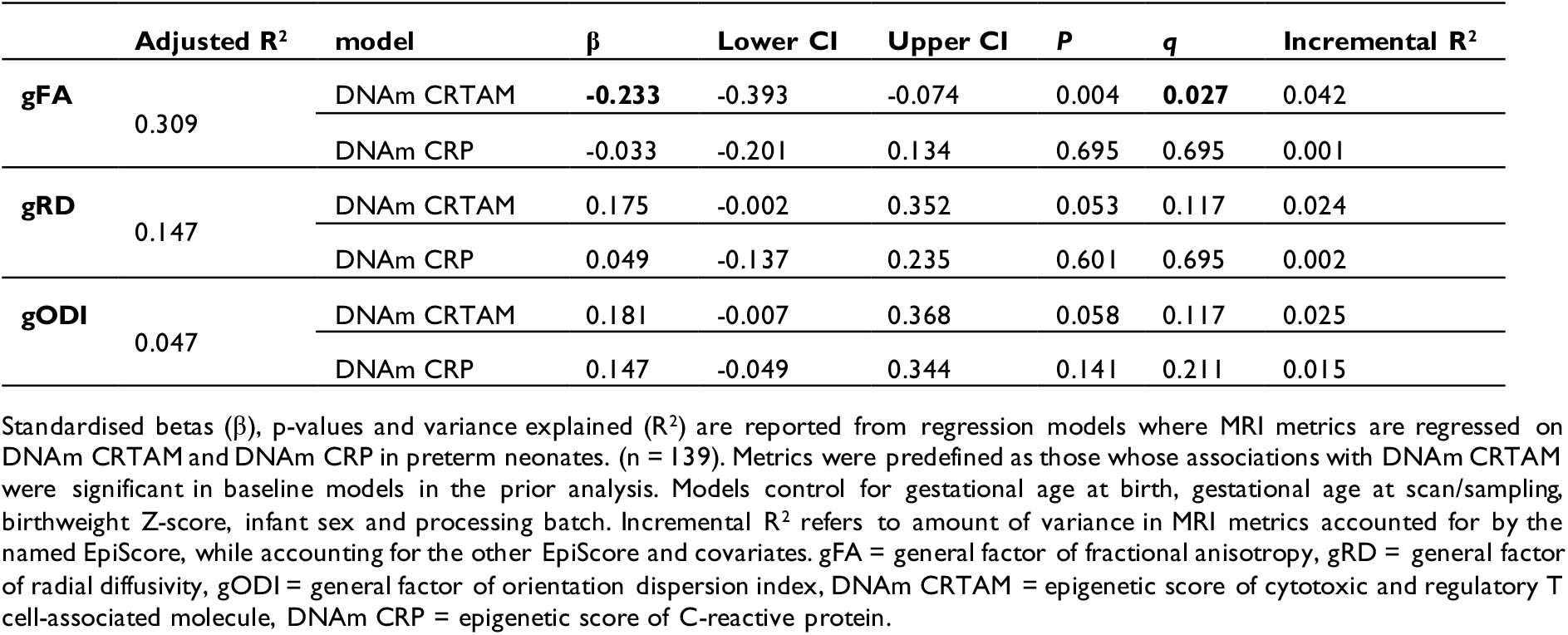
Associations of DNAm CRTAM with global metrics of neonatal white matter microstructure, accounting for contribution of DNAm CRP.

Despite numerically larger effect sizes for DNAm CRTAM compared to DNAm CRP, formal comparison using Williams’ test^82^ found no statistically significant differences in magnitude (*g*FA: t = −1.94, *P* = 0.054; *g*RD: t = 1.21, *P* = 0.23; *g*ODI: t = 0.35, *P* = 0.73). The comparison for *g*FA provided weak evidence for a larger association with DNAm CRTAM than DNAm CRP, although this was just at the statistical significance threshold.

### Unique contribution of DNAm CRTAM to variance in 5-year cognition beyond DNAm CRP

In preterm children with 5-year MSEL Early Learning Composite scores, DNAm CRTAM remained significantly associated with cognition after adjusting for DNAm CRP and other covariates (β = −0.332 [95% CI = −0.575, −0.089], *P* = 0.008, adjusted R^2^ = 17.1%, incremental R^2^ = 9.4%; Fig. 3). Notably, the coefficient for DNAm CRP was smaller and not statistically significant (β = −0.119, [95% CI = −0.362, 0.124], *P* = 0.33, incremental R^2^ = 1.2%). Together, these indicate that neonatal DNAm CRTAM accounts for more variance in 5-year cognition than DNAm CRP.

**Figure 3.**
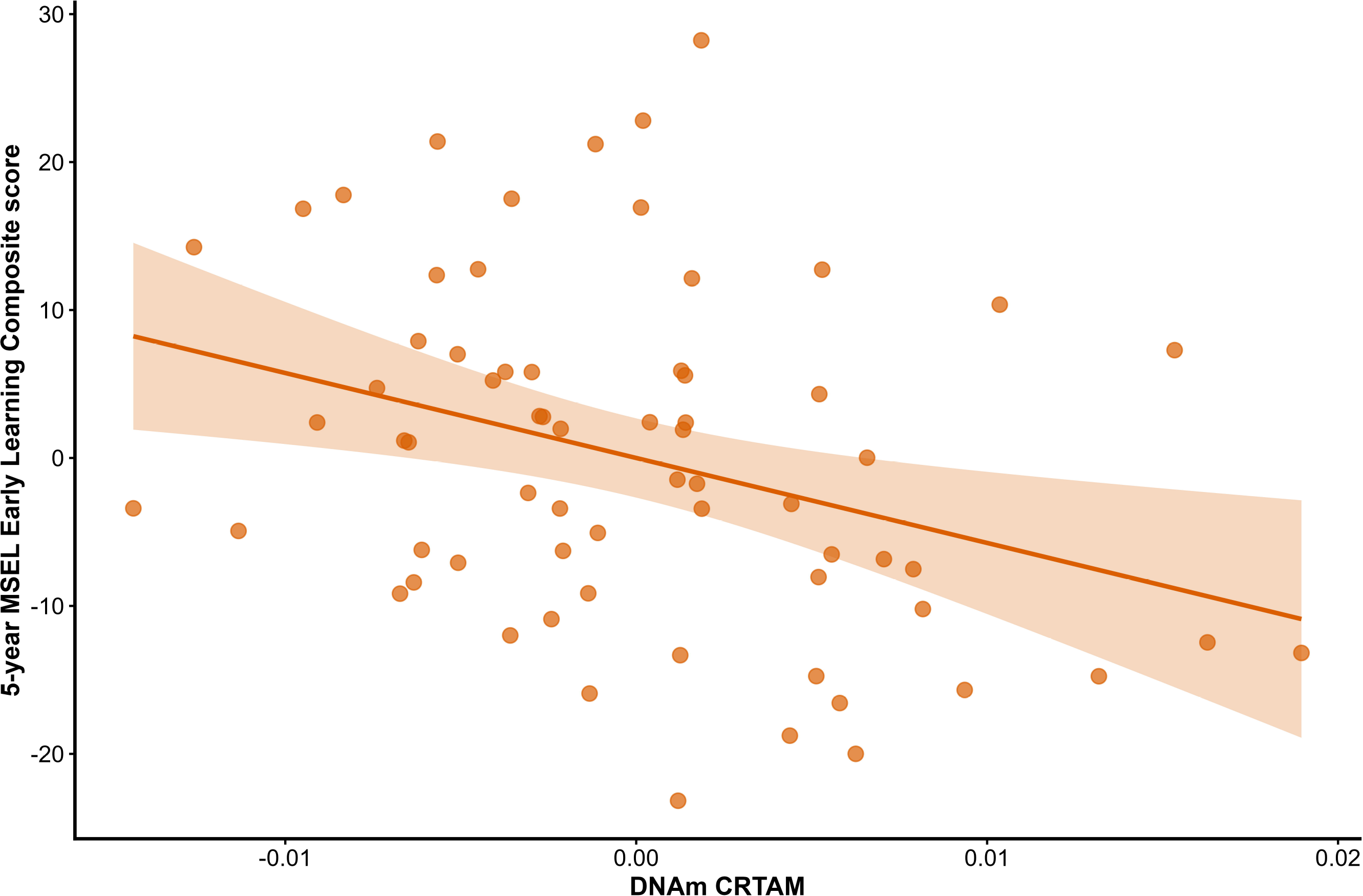
Partial regression plot showing the association between neonatal DNAm CRTAM and 5-year cognitive outcome independent of DNAm CRP. DNAm CRTAM and MSEL Early Learning Composite scores were residualised for DNAm CRP, gestational age at sampling, sex and DNAm processing batch. Points represent individual participants; line shows fitted association with 95% confidence interval. MSEL, Mullen Scales of Early Learning.

No interaction effect was observed between DNAm CRTAM and gestational age at birth on 5-year Early Learning Composite (product interaction term β = −0.036, *P* = 0.74), demonstrating consistent strength of association across the gestational age range.

## Discussion

We report the relevance of a novel immune-related, methylation-based, neonatal biomarker of the cytotoxic and regulatory T cell-associated molecule, DNAm CRTAM, to the EoP and later neurocognitive outcomes. Higher DNAm CRTAM, measured in saliva, is associated with MRI metrics of dysmature global white matter microstructure averaged across major tracts, consistent with EoP. Associations are robust to adjustment for inflammatory morbidities and a marker of non-specific inflammation, DNAm CRP. Similarly, associations of neonatal DNAm CRTAM with 5-year cognitive outcomes are independent of DNAm CRP. These findings suggest a potential role for CRTAM in EoP and functional relevance to later neurodevelopmental outcomes, with DNAm CRTAM reflecting an innate vulnerability of preterm infants that is not solely explained by inflammatory complications.

DNAm CRTAM is particularly interesting as a marker with both immune and neural relevance. The protein CRTAM is a promising marker for monitoring activation of cytotoxic and regulatory lymphoid cells. CRTAM^+^ T cells exhibit cytotoxic function, inducing marked expression of IFN-γ and other cytolytic genes.^83,84^ Interestingly, depletion of γδT cells, on which CRTAM is rapidly expressed following engagement of the T-cell receptor,^85^ protects mice from preterm white matter injury after hypoxic ischaemia.^86^ Moreover, elevated numbers of γδT cells are found in postmortem brains of preterm infants with white matter injury.^86,87^

Our main analyses revealed associations of DNAm CRTAM with diffusion MRI metrics of global white matter microstructure, namely *g*FA, *g*RD and *g*ODI, supporting the hypothesis that immune dysregulation and inflammation are linked to white matter alterations and microstructural disturbances in the preterm brain. These measures of poorer global white matter microstructure may reflect diffuse non-necrotic white matter injury common in nearly half of very preterm infants^88–90^ and associated with lower cognitive and academic outcomes.^90–92^

Specifically, DNAm CRTAM associates with lower FA and higher RD across the white matter, likely reflecting poorer axonal maturation and organisation and delayed myelination, respectively.^12,93,94^ ODI in white matter is less studied than DTI metrics in preterm infants at term-equivalent age but differences in ODI have been reported in white matter, where they may reflect altered axonal microstructure associated with EoP^14,95–98;^ and in very preterm children at 6 and 7 years of age, ODI is increased in major white matter tracts compared with term-born children and correlates with poorer IQ.^99,100^

Associations of DNAm CRTAM with *g*FA, *g*RD, *g*ODI and *g*MD were robust to adjustment for perinatal inflammatory morbidities in sensitivity analyses, similar to DNAm CRP.^17^ In addition, DNAm CRTAM consistently explained more incremental variance than DNAm CRP in diffusion MRI metrics. These two patterns are consistent with T-cell activation contributing to the pathogenesis of EoP through pathways independent of acute inflammation. Interestingly, we observed no interaction of DNAm CRTAM with gestational age at birth. It is possible that DNAm CRTAM reflects activation of an immune pathway linked to white matter microstructural variation through mechanisms that are not limited to the most premature infants, but are instead relevant across the preterm gestational age range studied. Notably, we did not observe associations of DNAm CRTAM with MRI volumetric features. This may be because diffuse white matter dysmaturation is the cardinal feature of EoP, with DNAm CRTAM potentially being more sensitive to microstructural differences.

The relative contributions of DNAm CRTAM and DNAm CRP to cognitive outcomes in preterm children revealed greater incremental variance explained by DNAm CRTAM (9.4% vs 1.2%), as well as significant associations not observed with DNAm CRP. Our findings corroborate those of Margara-Escudero *et al*.,^39^ who recently showed that CRTAM, and not CRP, partly contributed to a proteomic signature of global cognitive function in adults with metabolic morbidities. Reactome enrichment analysis revealed that CRTAM is involved in immunoregulatory interactions between lymphoid and non-lymphoid cells, essential for maintaining neuro-immune homeostasis.^39,101^ These results suggest a role of T-cell activation as a pathway linking preterm birth with brain dysmaturation and later cognition. As protein EpiScores are increasingly explored as tools for disease risk stratification, ^43,102–104^ DNAm CRTAM may represent a particularly informative marker for neurodevelopmental outcomes in preterm children, with potential predictive value beyond DNAm CRP. Importantly, the epigenetic proxy explained approximately 10% of variance in CRTAM protein levels in original test sets^102^. Other protein EpiScores consistently outperform paired, measured proteins in associations with several phenotypes, even when they explain a comparatively smaller proportion of variance in protein concentrations.^42,44,104–107^

Strengths of this study include deep phenotyping across multiple axes. We integrated methylation data with multimodal neonatal neuroimaging, clinically relevant exposures and childhood cognitive assessment. To our knowledge, this is the first study to investigate CRTAM in neurodevelopment. In particular, our use of salivary, methylation-based proxies of CRTAM and CRP offer a non-invasive, cost-effective and more stable alternative to serial venepuncture of proteins in neonates. Moreover, our choice of MRI features was based on established EoP characterisations that support inference about underlying biological processes and microstructural properties such as brain water content, myelination and axonopathy. We used the XTRACT protocol,^72,73^ which enabled us to derive microstructural measures of 42 major white matter tracts, achieving high brain coverage.

There are some limitations. First, the findings warrant replication studies in other cohorts, including those in low resource settings to assess generalisability; harmonisation of longitudinal data collection across modality types would support this goal. Second, the application of adult-trained EpiScores in developmental contexts with differing tissue types (e.g. blood versus saliva) and ancestries is a well-recognised challenge in the field^108,109^. However, growing evidence suggests that adult-trained EpiScores can be informative in paediatric samples^46,110,111^. In addition, moderate cross-tissue similarity has been demonstrated between blood-trained and saliva-trained CRP EpiScores (intra-class correlation coefficient = 0.58)^112^ and a blood-trained CRP EpiScore applied to saliva correlates similarly with serum CRP (r = 0.24-0.27).^55,113,114^ High concordance has been reported between salivary and blood DNAm patterns (r ∼0.97), as well as between salivary and brain tissue DNAm patterns (r ∼0.90), including among CpG sites not associated with methylation quantitative trait loci.^115,116^ While cross-tissue concordance may reflect shared or genetically influenced methylation patterns across tissues rather than necessarily indexing brain-specific methylation, these findings may support the utility of salivary-based EpiScores as accessible biomarkers in relation to neurocognitive phenotypes.^114^ Nonetheless, although DNAm CRP has been validated against serum CRP in cord blood,^54,56^ DNAm CRTAM has not yet been similarly validated against corresponding protein-level measures in a developmental cohort. Third, longitudinal DNAm and MRI at multiple time points would permit causal inference approaches to disentangle immune and neural functions of CRTAM in preterm brain dysmaturation, for example test whether alterations in white matter microstructure mediate the association between CRTAM EpiScore and brain or behavioural trajectories.

In conclusion, we observed relationships between a neonatal methylation-based proxy of CRTAM and the EoP and cognition at 5 years, above and beyond a proxy of CRP, highlighting activation of T-cell pathways as a potential contributor to preterm brain dysmaturation. The relevance of DNAm CRTAM to white matter dysmaturation-related phenotypes and its utility for risk stratification of neurodevelopmental outcomes should be further investigated.

## Data availability

Anonymised data included in these analyses are deposited in Edinburgh DataVault (https://doi.org/10.7488/e65499db-2263-4d3c-9335-55ae6d49af2b) and are available to researchers under the terms of the Theirworld Edinburgh Birth Cohort Data Access and Collaboration policy available at https://reproductive-health.ed.ac.uk/theirworld-edinburgh-birth-cohort-tebc/for-researchers/data-access-and-collaboration. The data are not part of an open repository due to the terms of the informed parent/guardian consent, which stipulates that the use of anonymised data is for studies of perinatal health that have been approved by regulatory bodies.

## Acknowledgements

The authors are grateful to the families of the Theirworld Edinburgh Birth Cohort who consented to take part in the study and to all the imaging research staff at the Edinburgh Imaging Facility for providing support with infant scanning. We thank Selina Abel, Melissa Thye and Jessica Dolan for substantial contributions to MSEL data collection.

## Funding

RS-B was supported by the Translational Neuroscience Programme at the University of Edinburgh, funded by Wellcome (218493/Z/19/Z). This work is supported by a UKRI Medical Research Council Programme Grant, MR/X003434/1 awarded to JPB. SRC was supported a Sir Henry Dale Fellowship jointly funded by the Wellcome Trust and the Royal Society (221890/Z/20/Z). REM was supported by UKRI Biotechnology and Biological Sciences Research Council (UKRI/BB/C001941/1) and Medical Research Council (UKRI3688) grants. The Theirworld Edinburgh Birth Cohort was initiated and is maintained by Theirworld (www.theirworld.org). The funders had no role in study design, data collection and analysis, decision to publish, or preparation of the manuscript.

For the purpose of open access, the authors have applied a CC BY public copyright licence to any Author Accepted Manuscript version arising from this submission.

## Competing interests

LM has received speaker and consultancy fees from Illumina. REM is a scientific advisor to the Epigenetic Clock Development Foundation and to Optima partners. All other authors declare that they have no competing interests.

## Supplementary material

Supplementary material is available online.

## Notes

### Author Declarations

The UK National Research Ethics Service, South East Scotland Research Ethics Committee (11/55/0061, 13/SS/0143) and NHS Lothian Research and Development (16/SS/0154) gave ethical approval for this work.

