## Supplementary material for "T-cell dysregulation is implicated in the encephalopathy of prematurity"

**Author affiliations**

This document includes:

- Supplementary Fig. 1. Raw tract-averaged values of each diffusion MRI metric across 42 brain white matter tracts.
- Supplementary Table 1. Tract loadings and explained variance for the first unrotated principal component (g-factor) for each diffusion MRI metric.
- Supplementary Fig 2. Scree plot showing percentage of variance explained and eigenvalues for the first ten principal components per diffusion MRI metric.
- Supplementary Fig. 3. Model diagnostics for association of DNAm CRTAM with general factor of fractional anisotropy (gFA).
- Supplementary Table 2. Associations of DNAm CRTAM with neonatal brain structure in preterm infants.
- Supplementary Table 3. Interaction effects of DNAm CRTAM and gestational age at birth on neonatal white matter microstructure in preterm infants.
- Supplementary Fig.4. Scatter plot showing correlation between DNAm CRTAM and DNAm CRP.
- Supplementary Table 4. Associations of DNAm CRTAM with global metrics of neonatal white matter microstructure, accounting for contribution of DNAm CRP.

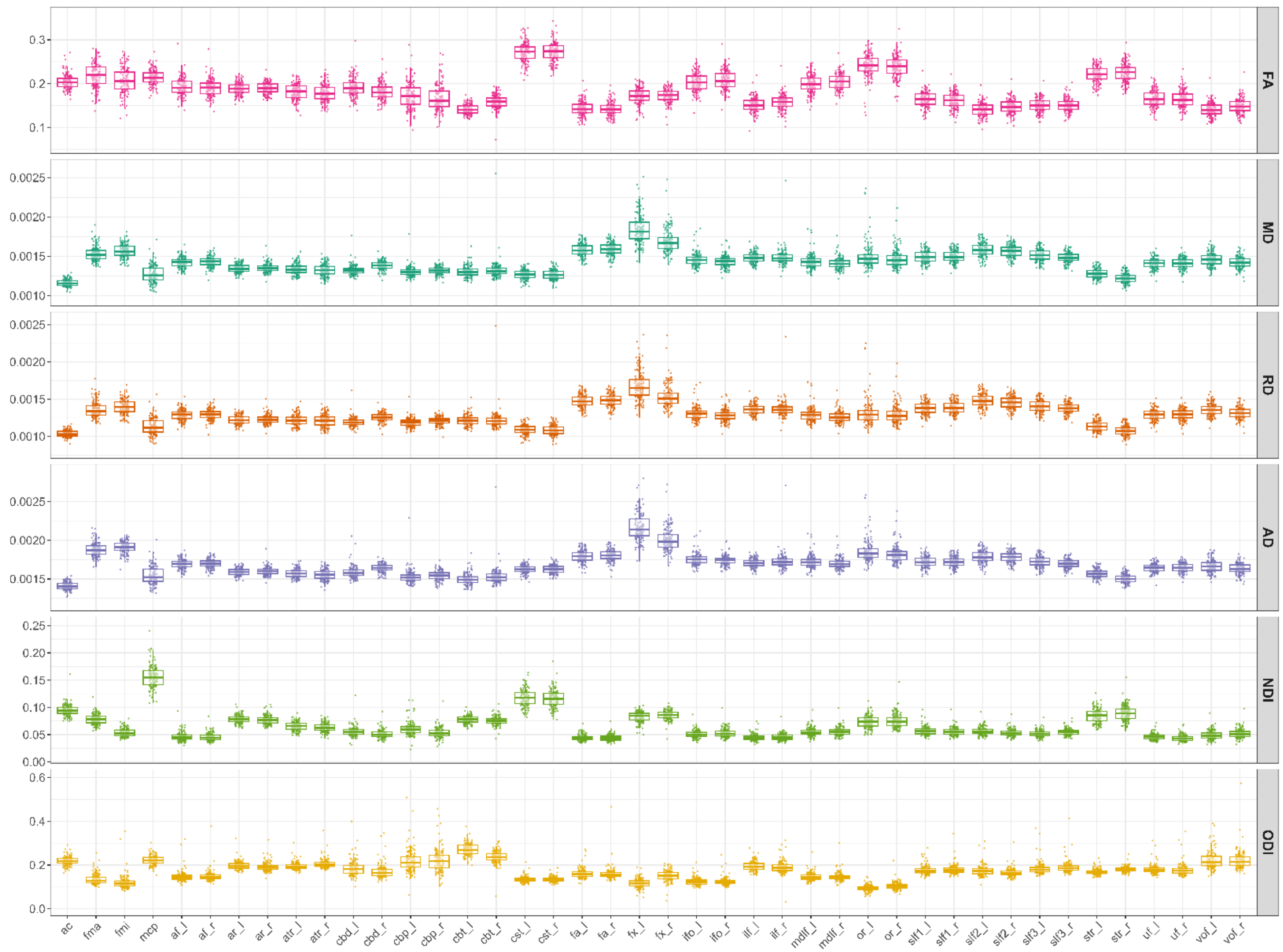


**Supplementary Fig. 1. Raw tract-averaged values of each diffusion MRI metric across 42 brain white matter tracts.** FA, fractional anisotropy; MD, mean diffusivity; RD, radial diffusivity; AD, axial diffusivity; NDI, neurite density index; ODI, orientation dispersion index; ac, anterior commissure; fma, forceps major (splenium of the corpus callosum); fmi, forceps minor (genu of the corpus callosum); mcp, middle cerebellar peduncle; af, arcuate fasciculus; ar, acoustic radiation; atr, anterior thalamic radiation; cbd, cingulum bundle: dorsal section; cbp, cingulum bundle: perigenual section; cbt, cingulum bundle: temporal section; cst, corticospinal tract; fa, frontal aslant tract; fx, fornix; ifo, inferior fronto-occipital fasciculus; ilf, inferior longitudinal fasciculus, mdlf, middle longitudinal fasciculus; or, optic radiation; slf1, slf2 and slf3, superior longitudinal fasciculus 1,2 and 3; str, superior thalamic radiation; uf, uncinate fasciculus; vof, vertical occipital fasciculus. '_l' and '_r' indicate left and right (bilateral) tracts.

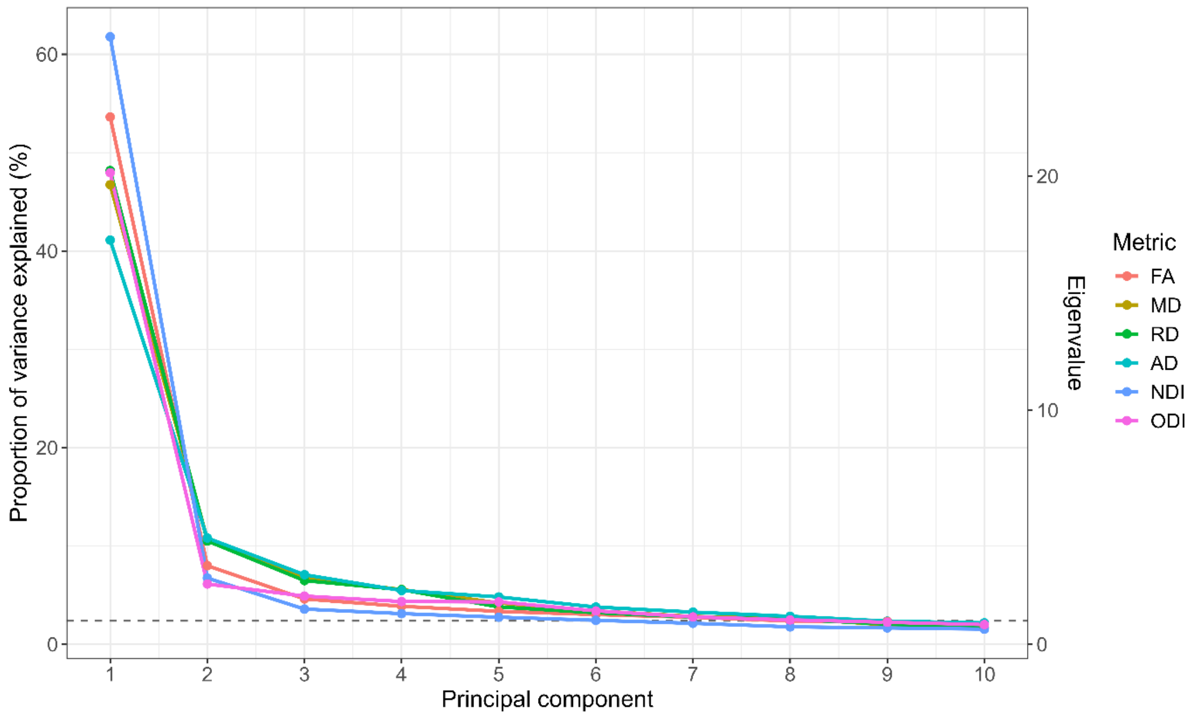


**Supplementary Fig. 2. Scree plot showing percentage of variance explained and eigenvalues for the first ten principal components per diffusion MRI metric.** FA, fractional anisotropy; MD, mean diffusivity; RD, radial diffusivity; AD, axial diffusivity; NDI, neurite density index; ODI, orientation dispersion index.

**Supplementary Table 1. Tract loadings and explained variance for the first unrotated principal component (g-factor) for each diffusion MRI metric.**

| tract | gFA | gMD | gRD | gAD | gNDI | gODI |
|---|---|---|---|---|---|---|
| ac | 0.356 | 0.329 | 0.317 | 0.355 | 0.689 | 0.331 |
| fma | 0.453 | 0.531 | 0.512 | 0.557 | 0.772 | 0.682 |
| fmi | 0.792 | 0.819 | 0.831 | 0.715 | 0.786 | 0.827 |
| mcp | 0.568 | 0.156 | 0.197 | 0.089 | 0.719 | 0.376 |
| af_l | 0.911 | 0.886 | 0.899 | 0.826 | 0.909 | 0.848 |
| af_r | 0.908 | 0.893 | 0.905 | 0.841 | 0.892 | 0.849 |
| ar_l | 0.631 | 0.634 | 0.626 | 0.618 | 0.791 | 0.57 |
| ar_r | 0.766 | 0.684 | 0.694 | 0.645 | 0.784 | 0.766 |
| atr_l | 0.722 | 0.798 | 0.8 | 0.762 | 0.793 | 0.719 |
| atr_r | 0.791 | 0.832 | 0.844 | 0.773 | 0.835 | 0.812 |
| cbd_l | 0.609 | 0.435 | 0.519 | 0.291 | 0.771 | 0.706 |
| cbd_r | 0.725 | 0.839 | 0.847 | 0.748 | 0.723 | 0.798 |
| cbp_l | 0.48 | 0.255 | 0.362 | 0.121 | 0.544 | 0.693 |
| cbp_r | 0.39 | 0.699 | 0.681 | 0.509 | 0.581 | 0.592 |
| cbt_l | 0.376 | 0.356 | 0.362 | 0.332 | 0.51 | 0.199 |
| cbt_r | 0.405 | 0.214 | 0.21 | 0.244 | 0.631 | 0.391 |
| cst_l | 0.738 | 0.733 | 0.729 | 0.71 | 0.852 | 0.521 |
| cst_r | 0.818 | 0.774 | 0.786 | 0.717 | 0.863 | 0.514 |
| fa_l | 0.829 | 0.804 | 0.807 | 0.785 | 0.769 | 0.77 |
| fa_r | 0.838 | 0.777 | 0.776 | 0.774 | 0.814 | 0.811 |
| fx_l | 0.265 | 0.235 | 0.216 | 0.291 | 0.15 | 0.13 |
| fx_r | 0.343 | 0.255 | 0.247 | 0.289 | 0.399 | 0.126 |
| ifo_l | 0.85 | 0.659 | 0.678 | 0.626 | 0.889 | 0.789 |
| ifo_r | 0.904 | 0.814 | 0.828 | 0.775 | 0.926 | 0.886 |
| ilf_l | 0.807 | 0.798 | 0.814 | 0.724 | 0.725 | 0.623 |
| ilf_r | 0.741 | 0.507 | 0.529 | 0.473 | 0.724 | 0.672 |
| mdlf_l | 0.753 | 0.581 | 0.591 | 0.566 | 0.87 | 0.673 |
| mdlf_r | 0.871 | 0.759 | 0.783 | 0.7 | 0.893 | 0.834 |
| or_l | 0.564 | 0.287 | 0.293 | 0.305 | 0.7 | 0.523 |
| or_r | 0.73 | 0.489 | 0.503 | 0.483 | 0.88 | 0.745 |
| slf1_l | 0.833 | 0.769 | 0.787 | 0.719 | 0.883 | 0.699 |
| slf1_r | 0.847 | 0.853 | 0.862 | 0.812 | 0.922 | 0.743 |
| slf2_l | 0.813 | 0.659 | 0.663 | 0.655 | 0.866 | 0.809 |
| slf2_r | 0.849 | 0.718 | 0.726 | 0.697 | 0.899 | 0.808 |
| slf3_l | 0.861 | 0.799 | 0.804 | 0.778 | 0.856 | 0.871 |
| slf3_r | 0.858 | 0.782 | 0.792 | 0.744 | 0.826 | 0.864 |
| str_l | 0.787 | 0.851 | 0.843 | 0.835 | 0.887 | 0.554 |
| str_r | 0.827 | 0.885 | 0.884 | 0.849 | 0.884 | 0.543 |
| uf_l | 0.849 | 0.813 | 0.83 | 0.75 | 0.785 | 0.836 |
| uf_r | 0.859 | 0.815 | 0.835 | 0.738 | 0.798 | 0.864 |
| vof_l | 0.758 | 0.699 | 0.737 | 0.61 | 0.807 | 0.712 |
| vof_r | 0.751 | 0.754 | 0.781 | 0.662 | 0.805 | 0.781 |
| Variance explained (%) | 53.632 | 46.734 | 48.175 | 41.112 | 61.783 | 47.962 |

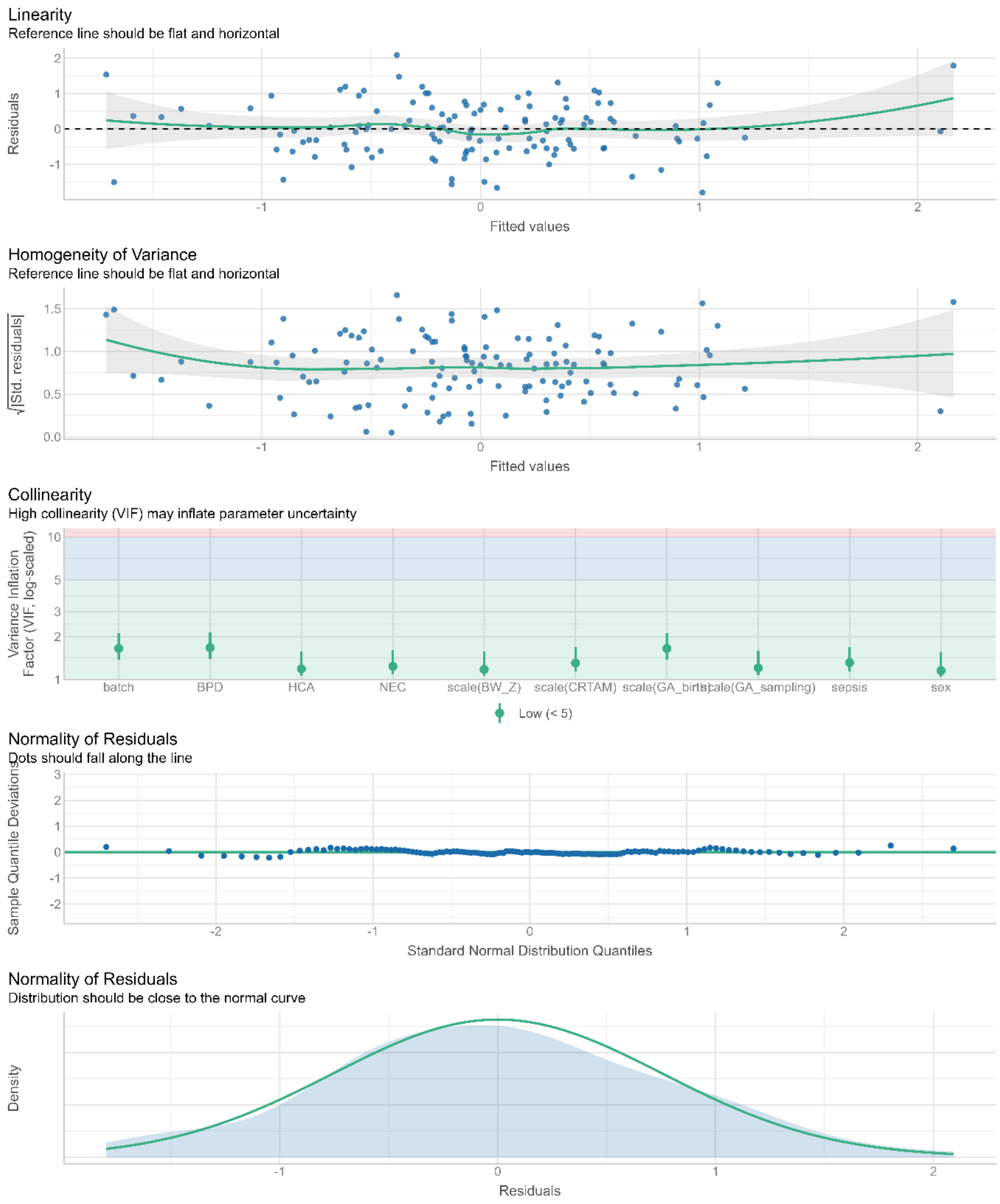


**Supplementary Fig. 3. Model diagnostics for association of DNAm CRTAM with general factor of fractional anisotropy (gFA).** Assumptions shown are tested for fully adjusted model (inflammation-associated model): gFA ~ DNAm CRTAM + gestational age at birth + gestational age at sampling/scan + infant sex + birthweight Z-score + DNAm processing batch + histologic chorioamnionitis + bronchopulmonary dysplasia + necrotising enterocolitis + sepsis.

**Supplementary Table 2. Associations of DNAm CRTAM with neonatal brain structure in preterm infants.**

| | model | Beta | Lower CI | Upper CI | p | Adjusted $R^2$ | n | Incremental $R^2$ | pFDR (q) |
|---|---|---|---|---|---|---|---|---|---|
| Cortical grey matter | Univariable model | 0.150 | -0.022 | 0.323 | 0.088 | 0.014 | 137 | NA | 0.323 |
| | Baseline model | 0.032 | -0.149 | 0.213 | 0.728 | 0.139 | 137 | 0.001 | 0.920 |
| | Inflammation-adjusted model | 0.025 | -0.163 | 0.212 | 0.796 | 0.111 | 126 | 0.000 | 0.898 |
| Deep grey matter | Univariable model | 0.041 | -0.133 | 0.216 | 0.642 | -0.006 | 137 | NA | 0.734 |
| | Baseline model | 0.010 | -0.186 | 0.206 | 0.920 | -0.016 | 137 | 0.000 | 0.920 |
| | Inflammation-adjusted model | -0.013 | -0.220 | 0.193 | 0.898 | 0.015 | 126 | 0.000 | 0.898 |
| White matter | Univariable model | -0.123 | -0.296 | 0.051 | 0.164 | 0.007 | 137 | NA | 0.323 |
| | Baseline model | -0.045 | -0.204 | 0.114 | 0.575 | 0.333 | 137 | 0.002 | 0.920 |
| | Inflammation-adjusted model | -0.066 | -0.229 | 0.097 | 0.424 | 0.346 | 126 | 0.003 | 0.830 |
| Hippocampi and amygdalae | Univariable model | -0.050 | -0.224 | 0.124 | 0.572 | -0.005 | 137 | NA | 0.734 |
| | Baseline model | -0.110 | -0.300 | 0.079 | 0.252 | 0.054 | 137 | 0.009 | 0.920 |
| | Inflammation-adjusted model | -0.117 | -0.317 | 0.084 | 0.251 | 0.064 | 126 | 0.010 | 0.830 |
| CSF | Univariable model | -0.009 | -0.184 | 0.165 | 0.917 | -0.007 | 137 | NA | 0.917 |
| | Baseline model | 0.048 | -0.146 | 0.242 | 0.624 | 0.008 | 137 | 0.002 | 0.920 |
| | Inflammation-adjusted model | 0.071 | -0.129 | 0.271 | 0.485 | 0.041 | 126 | 0.004 | 0.830 |
| Ventricles | Univariable model | -0.204 | -0.375 | -0.033 | 0.020 | 0.032 | 137 | NA | 0.160 |
| | Baseline model | -0.182 | -0.372 | 0.007 | 0.059 | 0.052 | 137 | 0.025 | 0.474 |
| | Inflammation-adjusted model | -0.144 | -0.333 | 0.045 | 0.134 | 0.041 | 126 | 0.017 | 0.830 |
| | Univariable model | 0.114 | -0.060 | 0.287 | 0.198 | 0.005 | 137 | NA | 0.323 |

| | | | | | | | | | |
|---|---|---|---|---|---|---|---|---|---|
| Brainstem | Baseline model | 0.024 | -0.170 | 0.218 | 0.808 | 0.011 | 137 | 0.000 | 0.920 |
| | Inflammation-adjusted model | 0.037 | -0.170 | 0.244 | 0.722 | -0.003 | 126 | 0.001 | 0.898 |
| Cerebellum | Univariable model | 0.112 | -0.061 | 0.286 | 0.202 | 0.005 | 137 | NA | 0.323 |
| | Baseline model | -0.011 | -0.195 | 0.174 | 0.910 | 0.103 | 137 | 0.000 | 0.920 |
| | Inflammation-adjusted model | -0.061 | -0.247 | 0.126 | 0.519 | 0.122 | 126 | 0.003 | 0.830 |
| gFA | Univariable model | -0.129 | -0.297 | 0.039 | 0.130 | 0.009 | 139 | NA | 0.391 |
| | Baseline model | **-0.239** | -0.396 | -0.083 | 0.003 | 0.313 | 139 | 0.046 | **0.018** |
| | Inflammation-adjusted model | **-0.252** | -0.408 | -0.096 | 0.002 | 0.374 | 128 | 0.051 | **0.011** |
| gMD | Univariable model | 0.075 | -0.093 | 0.244 | 0.379 | -0.002 | 139 | NA | 0.568 |
| | Baseline model | 0.165 | -0.013 | 0.342 | 0.068 | 0.117 | 139 | 0.022 | 0.102 |
| | Inflammation-adjusted model | **0.198** | 0.027 | 0.369 | 0.024 | 0.220 | 128 | 0.032 | **0.047** |
| gRD | Univariable model | 0.088 | -0.081 | 0.256 | 0.306 | 0.000 | 139 | NA | 0.568 |
| | Baseline model | 0.184 | 0.010 | 0.357 | 0.038 | 0.152 | 139 | 0.027 | 0.077 |
| | Inflammation-adjusted model | **0.215** | 0.046 | 0.383 | 0.013 | 0.251 | 128 | 0.038 | **0.039** |
| gAD | Univariable model | 0.036 | -0.133 | 0.205 | 0.673 | -0.006 | 139 | NA | 0.808 |
| | Baseline model | 0.104 | -0.080 | 0.289 | 0.265 | 0.043 | 139 | 0.009 | 0.265 |
| | Inflammation-adjusted model | 0.142 | -0.034 | 0.317 | 0.113 | 0.148 | 128 | 0.017 | 0.113 |
| gNDI | Univariable model | 0.002 | -0.167 | 0.171 | 0.985 | -0.007 | 139 | NA | 0.985 |
| | Baseline model | -0.103 | -0.244 | 0.038 | 0.151 | 0.439 | 139 | 0.008 | 0.182 |
| | Inflammation-adjusted model | -0.120 | -0.262 | 0.022 | 0.096 | 0.456 | 128 | 0.012 | 0.113 |
| gODI | Univariable model | 0.132 | -0.036 | 0.299 | 0.123 | 0.010 | 139 | NA | 0.391 |
| | Baseline model | 0.206 | 0.021 | 0.391 | 0.029 | 0.038 | 139 | 0.034 | 0.077 |

| | | | | | | | | | |
|---|---|---|---|---|---|---|---|---|---|
| | Inflammation-adjusted model | **0.201** | 0.018 | 0.384 | 0.032 | 0.023 | 128 | 0.036 | **0.048** |

Standardised betas and p-values are reported from regression models where MRI metrics are regressed on DNAm CRTAM. Bold text indicate statistically significant associations which survive FDR correction for multiple comparisons (q <0.05). Baseline models are controlled for gestational age at birth, gestational age at scan/sampling, infant sex, birthweight Z-score and DNAm processing batch. Inflammation-adjusted models additionally covary for all individual perinatal inflammatory morbidities simultaneously: histologic chorioamnionitis, sepsis, bronchopulmonary dysplasia and necrotising enterocolitis.

**Supplementary Table 3. Interaction effects of DNAm CRTAM and gestational age at birth on neonatal white matter microstructure in preterm infants.**

| | Model term | Beta | Lower CI | Upper CI | p | Adjusted R2 |
|---|---|---|---|---|---|---|
| gFA | DNAm CRTAM | -0.215 | -0.376 | -0.053 | 0.010 | 0.315 |
| | DNAm CRTAM x gestational age | 0.074 | -0.054 | 0.201 | 0.255 | |
| gRD | DNAm CRTAM | 0.152 | -0.028 | 0.331 | 0.097 | 0.157 |
| | DNAm CRTAM x gestational age | -0.095 | -0.236 | 0.047 | 0.187 | |
| gODI | DNAm CRTAM | 0.169 | -0.022 | 0.360 | 0.083 | 0.046 |
| | DNAm CRTAM x gestational age | -0.111 | -0.262 | 0.039 | 0.144 | |

Standardised betas and p-values are reported from regression models where MRI metrics are regressed on DNAm CRTAM (n = 139), including an interaction term (DNAm CRTAM x gestational age at birth). Models are controlled for gestational age at birth, gestational age at scan/sampling, infant sex, birthweight Z-score and DNAm processing batch.

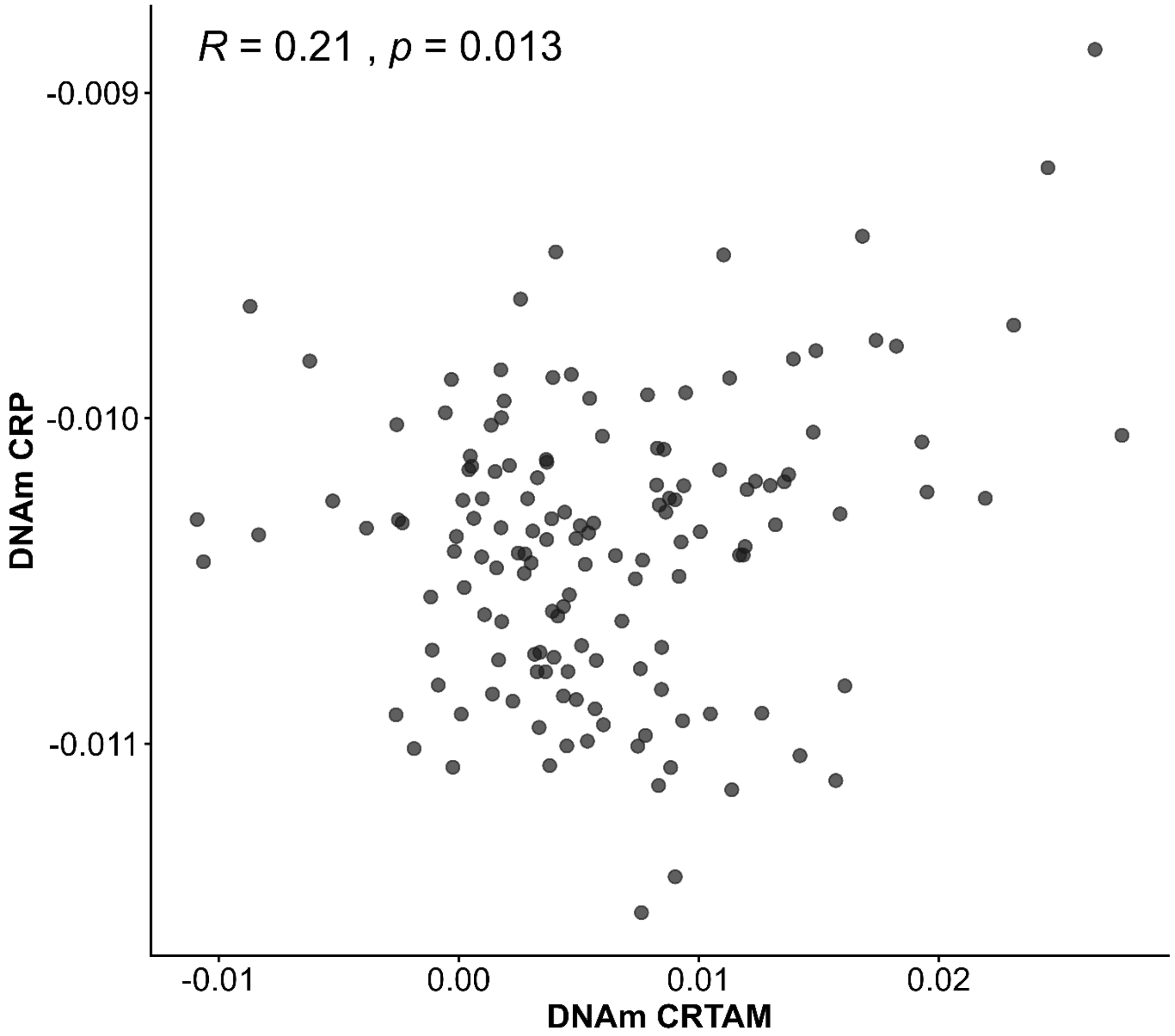


**Supplementary Fig. 4.** Scatter plot showing correlation between DNAm CRTAM and DNAm CRP.

**Supplementary Table 4. Associations of DNAm CRTAM with global metrics of neonatal white matter microstructure, accounting for contribution of DNAm CRP.**

| | **Adjusted $R^2$** | **model** | **β** | **Lower CI** | **Upper CI** | ***P*** | ***q*** | **Incremental $R^2$** |
|---|---|---|---|---|---|---|---|---|
| **Cortical grey matter** | 0.154 | CRTAM EpiScore | 0.018 | -0.162 | 0.197 | 0.847 | 0.934 | 0.000 |
| | | CRP EpiScore | 0.169 | -0.018 | 0.356 | 0.076 | 0.404 | 0.020 |
| **Deep grey matter** | -0.010 | CRTAM EpiScore | -0.002 | -0.198 | 0.195 | 0.986 | 0.986 | 0.000 |
| | | CRP EpiScore | 0.139 | -0.065 | 0.343 | 0.179 | 0.478 | 0.014 |
| **White matter** | 0.340 | CRTAM EpiScore | -0.034 | -0.193 | 0.125 | 0.672 | 0.895 | 0.001 |
| | | CRP EpiScore | -0.131 | -0.296 | 0.034 | 0.119 | 0.475 | 0.012 |
| **Hippocampi and amygdalae** | 0.059 | CRTAM EpiScore | -0.121 | -0.311 | 0.069 | 0.209 | 0.478 | 0.011 |
| | | CRP EpiScore | 0.127 | -0.069 | 0.324 | 0.203 | 0.478 | 0.011 |
| **CSF** | 0.008 | CRTAM EpiScore | 0.057 | -0.138 | 0.252 | 0.563 | 0.819 | 0.002 |
| | | CRP EpiScore | -0.104 | -0.306 | 0.098 | 0.309 | 0.593 | 0.008 |
| **Ventricles** | 0.049 | CRTAM EpiScore | -0.188 | -0.379 | 0.002 | 0.053 | 0.404 | 0.027 |
| | | CRP EpiScore | 0.071 | -0.127 | 0.269 | 0.478 | 0.766 | 0.004 |
| **Brainstem** | 0.010 | CRTAM EpiScore | 0.015 | -0.179 | 0.210 | 0.875 | 0.934 | 0.000 |
| | | CRP EpiScore | 0.099 | -0.103 | 0.301 | 0.334 | 0.593 | 0.007 |
| **Cerebellum** | 0.129 | CRTAM EpiScore | -0.028 | -0.211 | 0.154 | 0.760 | 0.934 | 0.001 |
| | | CRP EpiScore | 0.209 | 0.019 | 0.398 | 0.031 | 0.404 | 0.030 |
| **gFA** | 0.309 | CRTAM EpiScore | -0.233 | -0.393 | -0.074 | 0.004 | 0.053 | 0.042 |
| | | CRP EpiScore | -0.033 | -0.201 | 0.134 | 0.695 | 0.725 | 0.001 |
| **gMD** | 0.112 | CRTAM EpiScore | 0.157 | -0.024 | 0.337 | 0.089 | 0.249 | 0.019 |

| | | | | | | | | |
|---|---|---|---|---|---|---|---|---|
| | | CRP EpiScore | 0.046 | -0.144 | 0.236 | 0.631 | 0.725 | 0.001 |
| **gRD** | 0.147 | CRTAM EpiScore | 0.175 | -0.002 | 0.352 | 0.053 | 0.234 | 0.024 |
| | | CRP EpiScore | 0.049 | -0.137 | 0.235 | 0.601 | 0.725 | 0.002 |
| **gAD** | 0.037 | CRTAM EpiScore | 0.098 | -0.090 | 0.287 | 0.304 | 0.456 | 0.007 |
| | | CRP EpiScore | 0.035 | -0.162 | 0.233 | 0.725 | 0.725 | 0.001 |
| **gNDI** | 0.441 | CRTAM EpiScore | -0.119 | -0.262 | 0.025 | 0.104 | 0.249 | 0.011 |
| | | CRP EpiScore | 0.091 | -0.060 | 0.241 | 0.235 | 0.403 | 0.006 |
| **gODI** | 0.047 | CRTAM EpiScore | 0.181 | -0.007 | 0.368 | 0.058 | 0.234 | 0.025 |
| | | CRP EpiScore | 0.147 | -0.049 | 0.344 | 0.141 | 0.282 | 0.015 |

Standardised betas ($\beta$), p-values and variance explained ($R^2$) are reported from regression models where MRI metrics are regressed on DNAm CRTAM and DNAm CRP. (n = 139). Models control for gestational age at birth, gestational age at scan/sampling, birthweight Z-score, infant sex and processing batch. Incremental $R^2$ refers to amount of variance in MRI metrics accounted for by the named EpiScore, while accounting for the other EpiScore and covariates.
